# Findings from serological surveys (in August 2020) to assess the exposure of adult population to SARS Cov-2 infection in three cities of Odisha, India

**DOI:** 10.1101/2020.10.11.20210807

**Authors:** Jaya Singh Kshatri, Debdutta Bhattacharya, Srikanta Kanungo, Sidhartha Giri, Subrata Kumar Palo, Debaprasad Parai, Jyotirmayee Turuk, Asit Mansingh, Hariram Choudhary, Girish Chandra Dash, Niranjan Mishra, D.M. Satapathy, Sanjaya Ku Sahoo, Sanghamitra Pati

## Abstract

**Background:** There is always an uncertainty of epidemiological, serological infectivity and virulence of the emerging novel coronavirus. Antibody test can be used for assessing whether immunity has developed in the infected person after 5-7 days of illness and understand cumulative exposure levels to the infection, make inferences on the actual burden of infection, its geographical spread, effect on specific demographic/risk groups, gaps in testing and infection fatality rates.

**Objective:** To estimate and compare the sero-prevalence, hidden prevalence and determine the demographic risk factors associated with SARS-CoV-2 infection among adults in three largest cities of Odisha, India.

**Methodology:** This was a population based cross sectional serological survey carried out in August 2020 in the three largest cities of the state of Odisha. Sample size per city was estimated to be 1500 and participants were enrolled from the community using multi-stage random sampling from 25 clusters from each city. Data was collected using ODK based tools by household visits and 3-4 ml of blood samples were collected after informed consent. Samples were transported to testing lab where Serum was separated and tested for anti-SARS CoV-2 antibodies using automated CLIA platform. Statistical analysis was done using R-software packages.

**Results:** A total of 4146 participants from the 3 cities of Bhubaneswar (BBS), Berhampur (BAM) and Rourkela (RKL) participated. A total of 5635 households were approached and the average non response rate in the community was 17.4%. The gender weighted seroprevalence across the three cities was 20.78% (95% CI: 19.56%-22.05%). Seroprevalence was highest in BAM at 31.14% (95% CI: 28.69-33.66%) followed by 24.59% (95% CI: 22.39-26.88%) in RKL and 5.24% (95% CI: 4.10-6.58%) in BBS. While females reported a higher seroprevalence (22.8%) as compared to males (18.8%), there was no significant difference in seroprevalence across age groups. A majority of the seropositive participants were asymptomatic (93.87%). Among those who reported symptoms, the most common symptom was fever (68.89%) followed by cough (46.06%) and myalgia (32.67%). The case to infection ratio on the date of serosurvey was 1: 6.6 in BBS, 1:61 in BAM and 1:29.8 in RKL.

**Conclusion:** The study found a high seroprevalence against COVID-19 in urban Odisha as well as high numbers of asymptomatic infections.

## Background

The COVID19 pandemic has so far affected 216 countries and caused more than 32 million cases and about a million deaths worldwide.(1)India has the second largest number of cases at 5.9 million and has been reporting about a quarter of the daily global incident cases for some time now. (2) While the pandemic is at different stages across the country, there seems to be significant local differences in the progression within the states as well. The state of Odisha in eastern India contributes over 3% of the active case load but with less than 1% of the cumulative mortality in the country. (2) The pandemic in the state has till now been largely driven by urban clusters with major cities contributing the most to the caseload. (3) Large population size with high density, presence of slums, variable adherence to preventive measures and a sizeable migrant population seem to be the common characteristics driving the transmission of infection in these cities. These regions remain critical to the COVID-19 response of the state.

As with any novel respiratory infection, there is an uncertainty of epidemiological, serological, infectivity and virulence related information of SARS-CoV-2. (4)Testing strategies and capacities have been evolving but broadly, until now,it has been focused on the symptomatic and higher risk groups, which tend to overestimate the burden of the infection in the community due to biased denominator. (4–6) Additionally, the role of pre-symptomatic, asymptomatic and subclinical infection in disease transmission dynamics is also not well understood. (4)

While viral nucleic acid detection by real-time polymerase chain reaction (RT-PCR) test is considered the gold standard frontline test for clinical diagnosis of SARS-CoV-2, it is useful only when performed in the acute stage of infection.(7)Usually, antibody tests can be used for disease detection after 5–7 days of illness. IgM antibodies are evident in the blood for the first two months and IgG antibodies generally start appearing after two weeks of onset of infection and last for several months. (8– 10)Thus, although these tests are not useful for detecting acute infection, population based sero-epidemiological studies could be useful to understand cumulative exposure levels to the infection and make inferences on the actual burden of infection, its geographical spread, effect on specific demographic/risk groups, gaps in testing and infection fatality rates. (4,10)The information from such studies will also be helpful for designing and monitoring immunization programs, as and when a vaccine is available.(11)This evidence will inform the policy makers to plan and implement public health interventions for prevention and control of the pandemic. Thus the following sero survey is proposed to assess the extent of infection in the community and specific groups with the following objective:

### Objective

To estimate and compare the sero-prevalence, hidden prevalence and determine the demographic risk factors associated with SARS-CoV-2 infection among adults in three largest cities of Odisha, India.

### Methodology

This was a population based cross sectional serological survey carried out in August 2020 in the three largest cities of the state of Odisha in eastern India with a total population of over 2 million. The study population was randomly selected from the community members of the municipal wards from each of the city. Adults residing in the city since at least the past 3 months and who agreed to provide written informed consent for data and sample collection were included in the study. We excluded pregnant women, bed ridden patients and those with recognizable cognitive impairment. Minimum sample size per city was calculated to be 1437. This was done on the Open Epi ver3.0 software using the following formula-

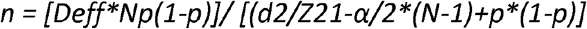

We assumed a sero-prevalence of 15%, which has been reported in urban regions of India during the same time period, relative precision of 20%, a design effect of 2.2 (calculated using a weak interclass correlation and a cluster size of 60), a finite population and a non-response rate of 20%. (12–14)This was rounded off to 1500 per city.

Multi-stage random sampling was used for recruiting participants. For every city, the municipal wards were treated as clusters and 25 wards were selected based on probability proportional to size. Residential street names in the ward were listed and the street from where the sampling began (as well as direction of sampling) in each cluster was selected by computerized simple random method. Households in the street were selected using systematic random sampling and one eligible individual was selected from each household using an age ordered matrix. Locked house and/or non-response were recorded and the sampling frame was shifted by one household to the immediate adjacent house in these cases. The sampling framework is provided below in figure-1.

**Figure-1.**
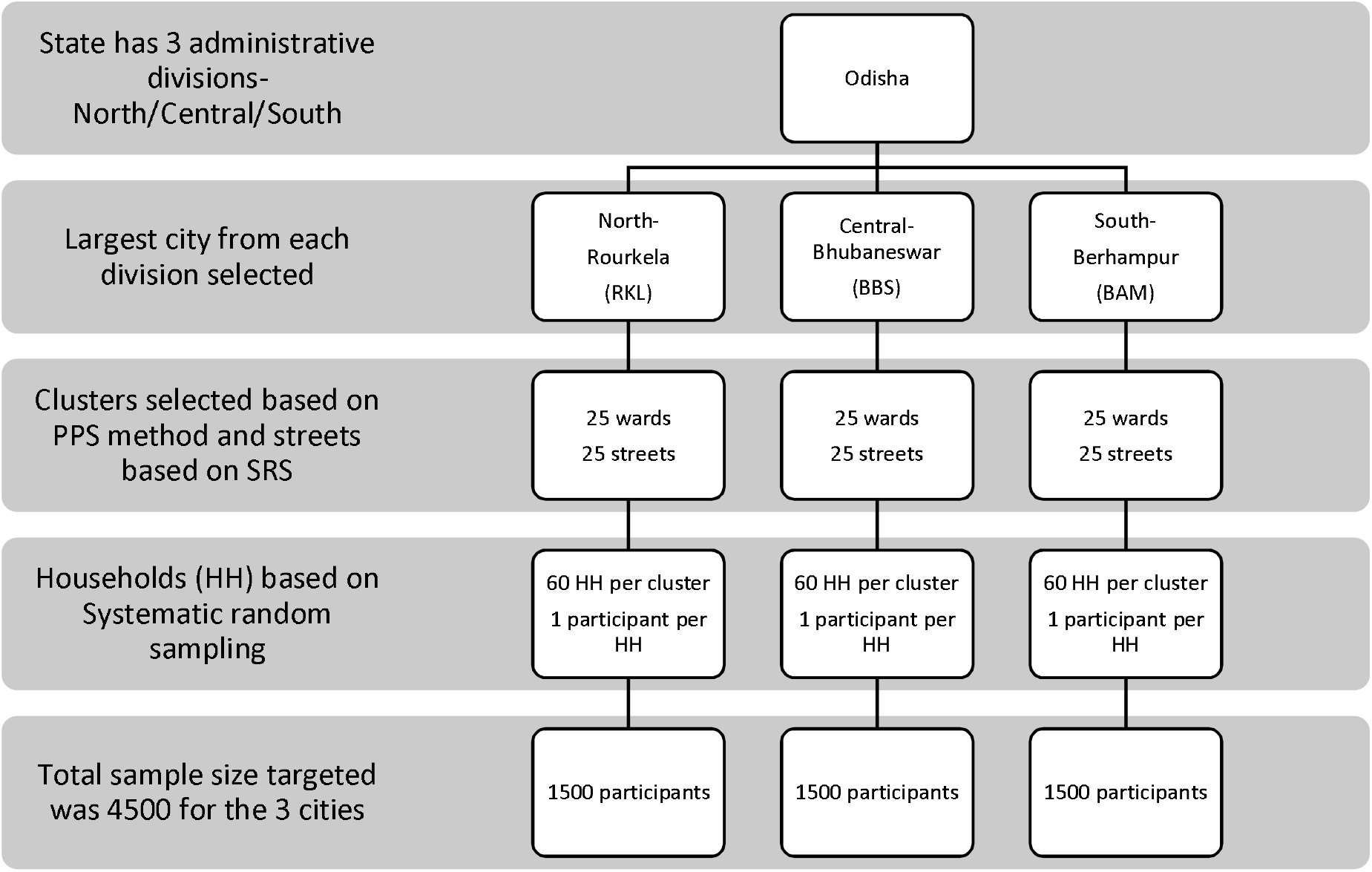
Sampling framework of the Multi-stage sampling design.

Data on the socio-demographic variables, exposure history with a confirmed (and/or suspected) case, symptom profile in the last 30 days, geographical location, travel and testing history were collected in a structured tool by trained field investigators who conducted participant interviews. An Open Data Kit based electronic data capture tool was used for this purpose. Following all aseptic precautions, 3-4 ml blood samples were collected in the field by trained phlebotomists by venepuncture and transferred to vacutainers. These were transported maintaining cold chain (2-8 C) to the serology laboratory at Indian Council of Medical Research-Regional Medical Research Centre in Bhubaneswar(ICMR-RMRC) for analysis. Additionally, secondary data on the daily number of antigen tests carried out, number of positives and deaths due to COVID-19 were obtained for the past 3 months from government sources directly.

Serum samples were subjected to detection in Roche Cobas e411 for the presence of IgG antibodies against COVID-19 using Electro-chemiluminescence immunoassay (ECLIA) based technique which is based on test principle of double-antigen sandwich assay and provides the result in 18 minutes. Elecsys® Anti-SARS-CoV-2 is an immunoassay for the in vitro qualitative detection of antibodies (including IgG) to SARS-CoV-2 in human serum and plasma. The assay uses a recombinant protein representing the nucleocapsid (N) antigen for the determination of antibodies against SARS-CoV-2. The test is intended as an aid in the determination of the immune reaction to SARS-CoV-2.

Testing procedures were followed as per the manufacturer’s instructions. Patient’s sample (20 μL) were incubated with a mix of biotinylated and ruthenylatednucleocapsid (N) antigen. Double-antigen sandwich immune complexes (DAGS) are formed in the presence of corresponding antibodies. After addition of streptavidin-coated microparticles, the DAGS complexes bind to the solid phase via interaction of biotin and streptavidin. After that the reagent mixture was transferred to the measuring cell, where the microparticles were magnetically captured onto the surface of the electrode. Unbound substances were subsequently removed. Electrochemiluminescence was then induced by applying a voltage and measured with a photomultiplier. The signal yield increased with the antibody titre. The value was expressed in Cut off Index (CoI) and a value of <1.0 was considered non-reactive and COI ≥1.0 was reactive.

The sero-prevalence of SARS-CoV-2 infection was estimated as proportion along with 95% confidence intervals and its distribution assessed across cities and demographic parameters. Gender weights were added in prevalence estimates in order to account for a higher non-response rate in females. The infection-to-case ratio and the infection fatality rate were calculated. Median time of seroconversion was assessed by a time dependent plot among those previously tested positive for SARS-CoV-2 by real time polymerase chain reaction (RT-qPCR). Temporal comparisons of the community seroprevalence estimates with the detected number of cumulative cases, active cases, recoveries and deaths was done. Heat maps for varying seroprevalence were built for each of the city’s wards. Statistical analyses were done using R (ver. 4.0.2) software packages and GIS analysis was done using QGIS (ver. 3.10).

Interviews were conducted ensuring privacy. All data was stored securely under the investigator’s responsibility, with a focus on ensuring the confidentiality of study participants. The final report and publications are based on aggregate data without any identifying information. A database with electronic tracking, password-restricted access and audit trail, with time and date stamps on data entry and edits, was used for quality control. State Health Department and city administration authorities were actively engaged to ensure smooth operationalization and also to reduce any stigma among the residents. Written informed consent for participation was obtained and a participant information sheet was provided to each household. Approval for the protocol was obtained from the ICMR RMRC Institutional Human Ethics Committee and the State Health and research ethics committee. The study methods, analyses and reporting have been informed by the WHO Unity protocols and ICMR National Serosurvey protocol in India.(15,16)

## Results

The study was conducted among 4146 participants from the 3 cities of Bhubaneswar (BBS), Berhampur (BAM) and Rourkela (RKL). A total of 5635 households were approached and the average non response rate in the community was 17.4%, which was similar across the three cities. The study flow diagram is provided below in figure-2.

**Figure-2.**
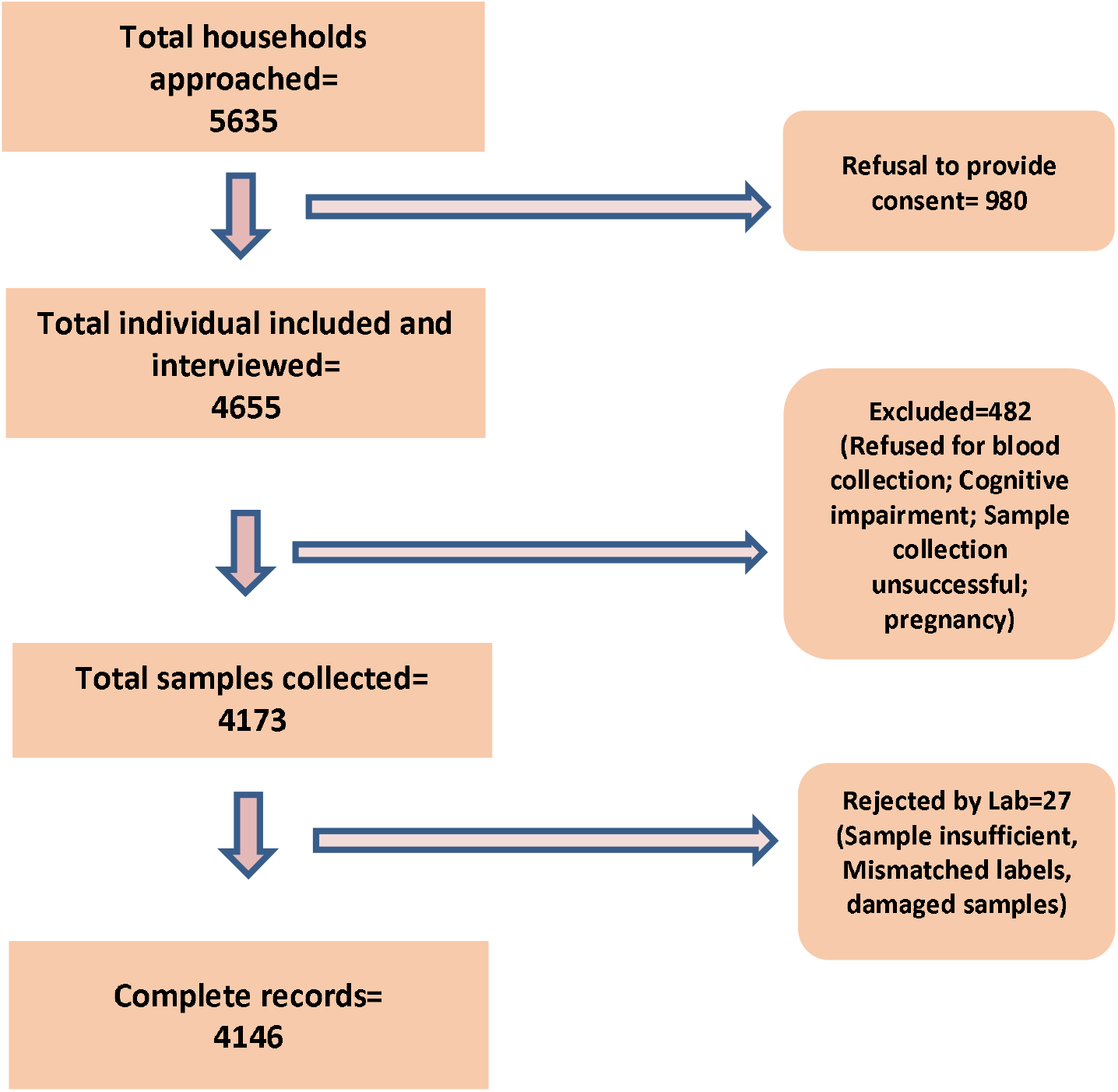
Study Flow diagram of the sample collection process.

Among the study participants, 1553 were females and the rest males. The non-response rates were significantly higher among females (27.6%) as compared to males (12.4%). The mean age of the study participants was 44.20 (± 14.2) years.

The gender weighted seroprevalence across the three cities was 20.78% (95% CI: 19.56%-22.05%). This was highest in BAM at 31.14% (95% CI: 28.69%-33.66%) followed by 24.59% (95% CI: 22.39%-26.88%) in RKL and 5.24% (95% CI: 4.10%-6.58%) in BBS. While females reported a higher seroprevalence (22.8%) as compared to males (18.8%), there was no significant difference in seroprevalence across age groups.

The demographic characteristics of the study population and the distribution of seroprevalence is provided in table-1 below.

**Table-1.** Demographic characteristics of the study population and distribution of seroprevalnce

| Table-1. Demographic characteristics of the study population and distribution of seroprevalence |  |  |  |  |  |  |  |  |  |  |
| --- | --- | --- | --- | --- | --- | --- | --- | --- | --- | --- |
| Characteristic |  | City |  |  |  |  |  | Total |  | χ<sup>2</sup><br>(p-value) |
|  |  | BBS |  | BAM |  | RKL |  |  |  |  |
|  |  | N= | Positive=<br>(%, 95% CI) | N= | Positive=<br>(%, 95% CI) | N= | Positive=<br>(%, 95% CI) | N= | Positive=<br>(%, 95% CI) |  |
| Age Group | 18-29 years | 276 | 16<br>(5.79%, 3.34-9.24%) | 177 | 57<br>(32.20%, 25.39-39.62%) | 256 | 58<br>(22.65%, 17.67-28.28%) | 709 | 131<br>(18.47%, 15.68-21.53%) | 8.318<br>(.081) |
|  | 30-39 years | 298 | 23<br>(7.71%, 4.95-11.35%) | 251 | 90<br>(35.85%, 29.92-42.12%) | 344 | 82<br>(23.83%, 19.43-28.69%) | 893 | 195<br>(21.83%, 19.16-24.69%) |  |
|  | 40-49 years | 311 | 08<br>(2.57%, 1.11-5.00%) | 364 | 114<br>(31.31%, 26.58-36.35%) | 351 | 80<br>(22.79%, 18.50-27.54%) | 1026 | 202<br>(19.68%, 17.29-22.25%) |  |
|  | 50-59 years | 258 | 09<br>(3.48%, 1.60-6.51%) | 271 | 77<br>(28.41%, 23.12-34.18%) | 287 | 65<br>(22.64%, 17.93-27.93%) | 816 | 151<br>(18.50%, 15.89-21.34%) |  |
|  | ≥ 60 years | 176 | 13<br>(7.38%, 3.99-12.29%) | 312 | 89<br>(28.52%, 23.58-33.88%) | 214 | 61<br>(28.50%, 22.22-35.05%) | 702 | 163<br>(23.21%, 20.14-26.52%) |  |
| Gender | Male | 891 | 46<br>(5.16%, 3.80-6.82%) | 847 | 260<br>(30.69%, 27.60-33.92%) | 855 | 182<br>(21.28%, 18.58-24.18%) | 2593 | 488<br>(18.81%, 17.33-20.37%) | 9.481<br>($<0.01$) |
|  | Female | 428 | 23<br>(5.37%, 3.43-7.95%) | 597 | 167<br>(27.97%, 24.40-31.76%) | 597 | 164<br>(27.47%, 23.92-31.24%) | 1553 | 354<br>(22.79%, 20.73-24.96%) |  |
|  | Others | 0 | 0 | 0 | 0 | 0 | 0 | 0 | 0 |  |
| Occupation | Manufacturing | 62 | 6<br>(9.67%, 3.63-19.88%) | 120 | 47<br>(39.16%, 30.38-48.49%) | 66 | 17<br>(25.75%, 15.77-38.00%) | 248 | 70<br>(28.22%, 22.71-34.26%) | 62.565<br>($<0.01$) |
|  | Public Service | 200 | 11<br>(5.5%, 2.77-9.62%) | 133 | 34<br>(25.56%, 18.40-33.84%) | 267 | 44<br>(16.47%, 12.23-21.48%) | 600 | 89<br>(14.83%, 12.08-17.93%) |  |
|  | Private Service | 295 | 7<br>(2.37%, 0.96-4.83%) | 161 | 41<br>(25.46%, 18.93-32.92%) | 226 | 40<br>(17.69%, 12.95-23.31%) | 682 | 88<br>(12.90%, 10.48-15.65%) |  |
|  | Self employed | 206 | 11<br>(5.33%, 2.69-9.35%) | 295 | 105<br>(35.59%, 30.12-41.34%) | 161 | 50<br>(31.05%, 24.00-38.81%) | 662 | 166<br>(25.07%, 21.81-28.55%) |  |
|  | Homemaker | 251 | 10<br>(3.98%, 1.92-7.20%) | 303 | 90<br>(29.70%, 24.61-35.19%) | 377 | 112<br>(29.70%, 25.13-34.60%) | 931 | 212<br>(22.77%, 20.11-25.60%) |  |
|  | Student | 73 | 3<br>(4.10%, 0.85-11.74%) | 58 | 18<br>(31.03%, 19.53-44.54%) | 76 | 16<br>(21.05%, 12.53-31.92%) | 207 | 37<br>(17.87%, 12.90-23.78%) |  |
|  | Unemployed | 101 | 8<br>(7.92%, 3.48-15.01%) | 144 | 38<br>(26.38%, 19.40-34.37%) | 129 | 39<br>(30.23%, 22.46-38.93%) | 374 | 85<br>(22.72%, 18.57-27.31%) |  |
|  | Others/Retired | 131 | 13<br>(9.92%, 5.39-16.37%) | 161 | 54<br>(33.54%, 26.30-41.39%) | 150 | 28<br>(18.66%, 12.77-25.83%) | 442 | 95<br>(21.49%, 17.75-25.62%) |  |
| Household Size | ≤ 2 | 239 | 10<br>(4.18%, 2.02-7.55%) | 228 | 50<br>(21.92%, 16.73-27.86%) | 185 | 38<br>(20.54%, 14.96-27.08%) | 652 | 98<br>(15.03%, 12.37-18.00%) | 37.457<br>($<0.01$) |
|  | 3-4 | 674 | 32<br>(4.74%, 3.26-6.63%) | 669 | 197<br>(29.44%, 26.01-33.06%) | 722 | 155<br>(21.46%, 18.52-24.64%) | 2065 | 384<br>(18.59%, 16.93-20.34%) |  |
|  | 5-6 | 307 | 18<br>(5.86%, 3.51-9.10%) | 339 | 129<br>(38.05%, 32.86-43.45%) | 375 | 102<br>(27.20%, 22.75-32.00%) | 1021 | 249<br>(24.38%, 21.78-27.14%) |  |
|  | >6 | 99 | 9<br>(9.09%, 4.24-16.55%) | 139 | 51<br>(36.69%, 28.68-45.27%) | 170 | 51<br>(30.00%, 23.22-37.49%) | 408 | 111<br>(27.20%, 22.94-31.08%) |  |
| Travel history | Yes | 74 | 1<br>(1.35%, 0.03-7.30%) | 22 | 6<br>(27.27%, 10.72-50.22%) | 26 | 4<br>(15.38%, 4.35-34.86%) | 122 | 11<br>(9.01%, 4.58-15.56%) | 9.904<br>($<0.01$) |
|  | No | 1245 | 68<br>(5.46%, 4.26-6.87%) | 1353 | 421<br>(31.11%, 28.65-33.65%) | 1426 | 342<br>(23.98%, 21.78-26.28%) | 4024 | 831<br>(20.65%, 19.40-21.93%) |  |
| Total |  | 1319 | 69<br>(5.23%; 4.09-6.47%) | 1375 | 427<br>(31.05%; 28.61-33.57%) | 1452 | 346<br>(23.82%; 21.62-26.10%) | 4146 | 842<br>(20.30%; 19.0-21.56%) | - |

Among the study population, 6.12% had developed symptoms suggestive of COVID-19 in the past 30 days and 9.98% had been tested by PCR for COVID-19. A majority of the seropositive participants were asymptomatic (93.87%). Among those who reported symptoms, the most common symptom was fever (68.89%) followed by cough (46.06%) and myalgia (32.67%). The distribution of seroprevalence according to symptoms and testing is given below in table-2.

**Table 2.** Testing status and symptom profile of the study participants

| Table-2. Testing status and symptom profile of the study participants |  |  |  |  |
| --- | --- | --- | --- | --- |
| Testing/Symptom profile | | N= | Seropositive<br>(%, 95% CI) | $\chi^2$<br>(p-value) |
| Tested by PCR | Yes | 414 | 90<br>(21.73%, 17.85-26.02%) | 0.581<br>(0.446) |
|  | No | 3732 | 752<br>(20.15%, 18.87-21.47%) |  |
| PCR Result | Positive | 29 | 23<br>(79.31%, 60.3%-92.0%) | 60.752<br>($<0.01$) |
|  | Negative | 385 | 67<br>(17.40%, 13.7%-21.6%) |  |
| Symptoms of self<br>(last 30 days) | Yes | 254 | 80<br>(31.49%, 25.83-37.59%) | 20.924<br>($<0.01$) |
|  | No | 3892 | 762<br>(19.57%, 18.34-20.86%) |  |
| Symptoms in Household members<br>(last 30 days) | Yes | 51 | 11<br>(21.56%, 11.29-35.32%) | 0.051<br>(0.822) |
|  | No | 4095 | 831<br>(20.29%, 19.07-21.55%) |  |
| Symptom profile | Fever | 175 | 64<br>(36.57%, 29.44-44.17%) | 29.85<br>($<0.01$) |
| | Cough | 117 | 37<br>(31.62%, 23.33-40.86%) | 9.525<br>($<0.01$) |
|  | Shortness of breath/ difficulty<br>in breathing | 74 | 12<br>(16.21%, 8.66-26.61%) | .586<br>(0.444) |
|  | Myalgia | 109 | 28<br>(25.68%, 17.79-34.94%) | 2.002<br>(0.157) |
|  | Headache | 83 | 24<br>(28.91%, 19.48-39.90%) | 3.877<br>(0.049) |
| | Diarrhea | 12 | 7<br>(58.33%, 27.66-84.83%) | 10.752<br>($<0.01$) |
|  | Sore throat | 34 | 12<br>(35.29%, 19.74-53.51%) | 4.75<br>(0.029) |
|  | Anosmia/loss of taste | 12 | 3<br>(25%, 5.48-57.18%) | 0.164<br>(0.686) |

Among those who had seroconversion and also had been tested positive by PCR, the median duration between both was 31 days.

The cumulative number of cases detected as on 1^st^ September were 11641 in BBS, 3277 in BAM and 5362 in RKL. The association between time trends of the progression of the daily new cases and cumulative cases and the time point of seroprevalence estimates is given in figure-3a and 3b below.

**Figure-3a.**
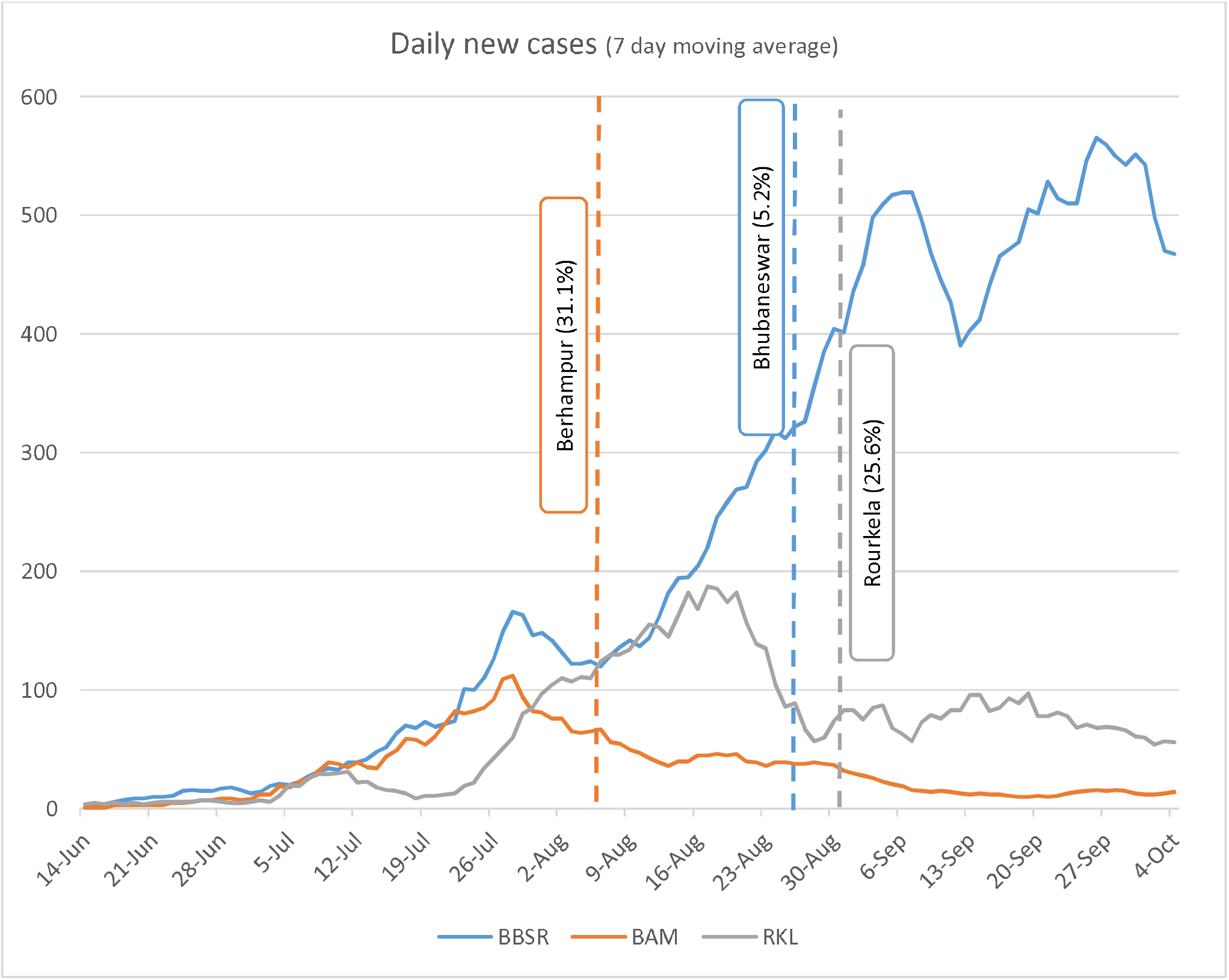
7 day moving average of new cases detected in three cities and point of seroprevalence.

**Figure-3b.**
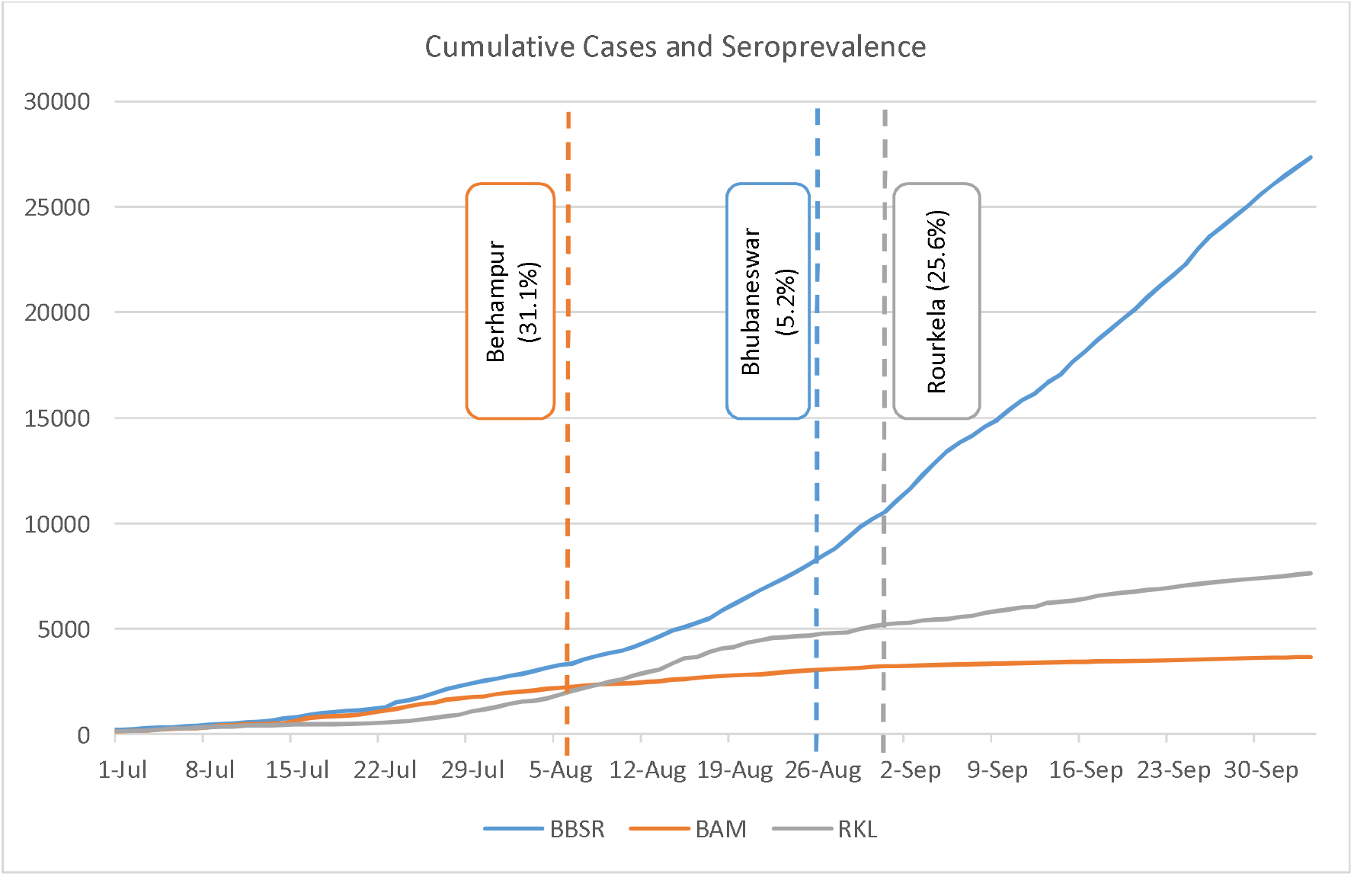
Cumulative cases detected in the three cities and point of seroprevalence estimates.

The case to infection ratio on the date of serosurveywas 1: 6.6 in BBS, 1:61 in BAM and 1:29.8 in RKL.The heat maps for the geographical distribution of the seroprevalence across the three cities is given in figure-4.

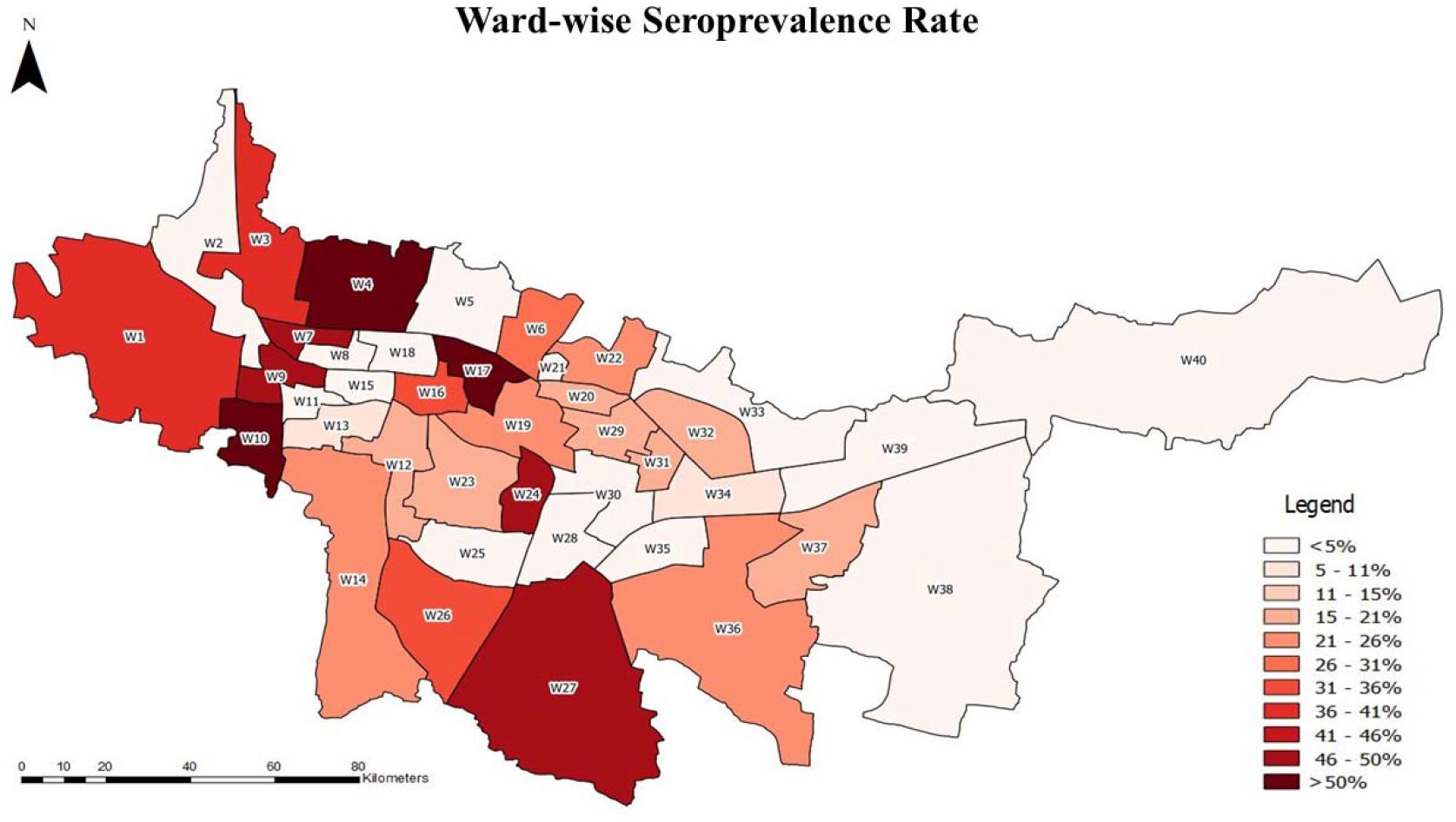
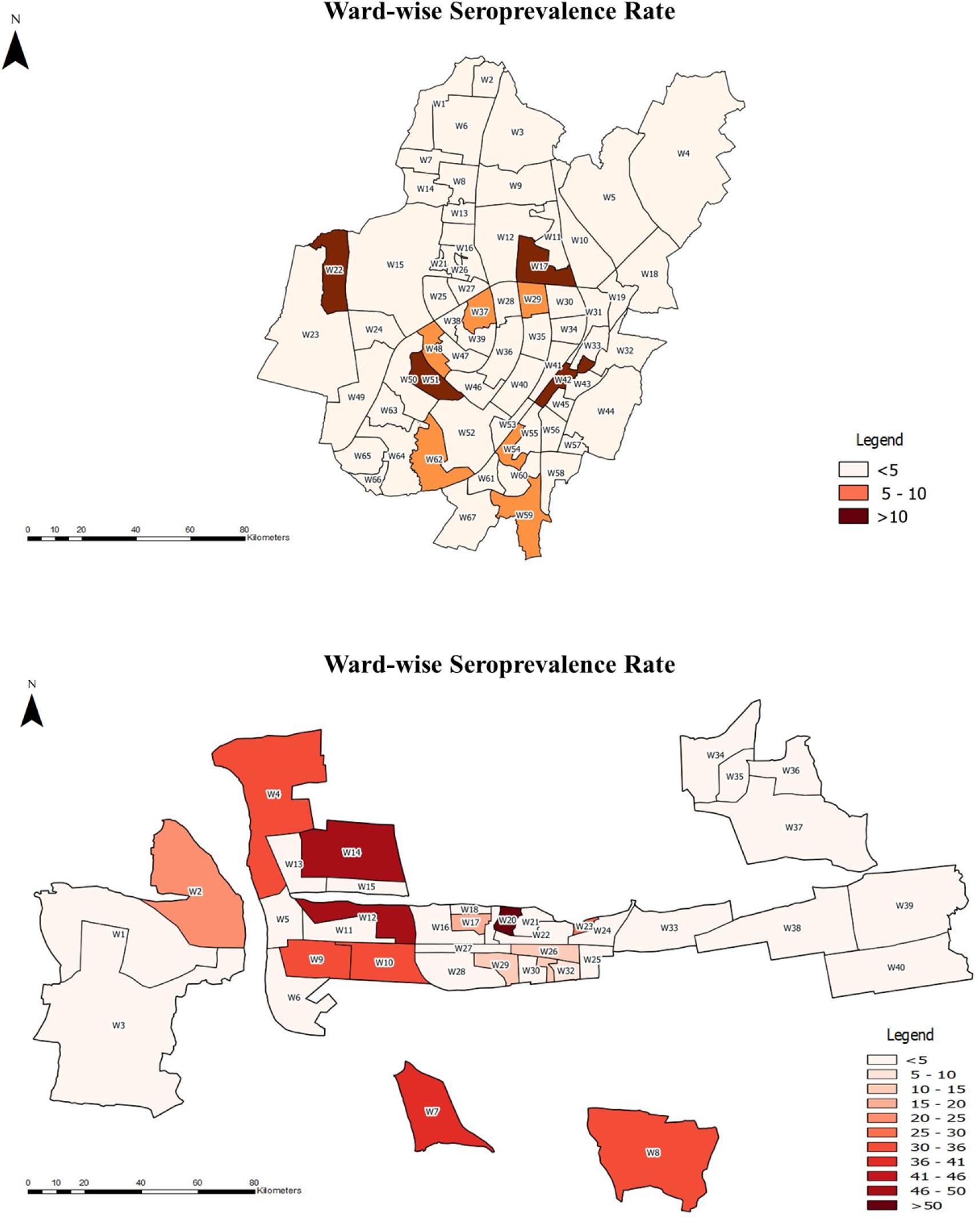

## Discussion

This COVID-19 serosurvey involving more than 4000 participants from the three largest cities of Odisha found an overall seroprevalence of 20.78%, although there was wide variation in seroprevalence between the cities. The study included only adult population. As educational institutions remain closed and lockdowns of various degrees have been imposed in the study setting since the pandemic began, the likelihood of adults being the source of household infection is high.

Although molecular tests are being used for diagnosis of active symptomatic and asymptomatic cases of COVID-19, antibody based tests can provide a more robust and comprehensive knowledge about the actual spread of infection in the community. (17)Seroprevalence studies on COVID-19 have been reported globally to assess the spread of infection either in the general population or focussing on certain high risk groups. Seroprevalence studies performed on health care workers across countries such as Germany, Belgium, United Kingdom, Malawi, and Italy, have reported wide variation in seropositivity ranging from 1.6% in Germany to 15.6% in Pakistan.(18–23) However, such studies on population involved in high risk professions, though useful in informing trends of infection in such occupations, do not provide information about exposure at the population level and inferences possible are limited.

Globally, there have been few reports on seroprevalence against COVID-19 in the community. An early study on seroprevalence from Lombardy, Italy, involving 390 blood donors showed that 23% of the donors were positive for anti-COVID-19 neutralizing antibodies. (24)A community-based study from British Columbia in Canada, involving serial cross-sectional sampling, reported a seroprevalence of only 0.28% in March 2020, and 0.55% in May 2020. (25)Another community based weekly serosurvey conducted in April 2020 in Geneva, Switzerland, reported a seroprevalence of 3.1%, 6.1%, and 9.7% during the first, second, and third week respectively, with a significantly higher seroprevalence in less than 50 years age group. (26)Such studies involving periodic sampling are extremely valuable in monitoring the spread of the infection in certain areas, and understanding the dynamics of community transmission as well as residual susceptibility. (25)In China, the seroprevalence in Guangzhou and Wuhan, the epicentre of the COVID-19 pandemic, was reported to be 0.6% and 2.1% respectively till April 2020. (27)During the same month, a study from Santa Clara County, California, reported a seroprevalence of 2.8% (95% CI 1.3-4.7%). (28)These studies across countries indicate that the actual spread of COVID-19 infection in the community was much higher than that reported by detection of active cases using molecular methods. Our study also presents evidence to support this case.

Few studies on seroepidemiology of COVID-19 have been reported from Asia till date. A national serosurvey from India conducted during May-June, 2020, which included 28000 individuals from 70 districts of 21 Indian states, reported the seroprevalence to be approximately 0.73% (95% CI: 0.34-1.13). (29)This indicated that the cumulative COVID-19 infections in India was approximately 6.46 million by the beginning of May 2020. Compared to our study, the low seroprevalence reported in the national serosurvey may be due to the difference in study period (May-June in the national serosurvey compared to August in our study). While our study showed females to be more infected, the national serosurvey found males to have significantly higher seroprevalence. The infection to case ratio (ICR) in this national serosurvey varied between 81.6 and 130.1 with May 11 and May 3, 2020, as reference points for reported cases. (29)This is higher than that reported in our study (ranging from 6.6 in Bhubaneswar to 61 in Berhampur). The steady increase in testing and subsequent improvement in case identification may be the reason for this difference. Similar to our study, the national serosurvey also reported a higher seropositivity rate in occupations with high risk of exposure to potentially infected persons. (29)Very few other countries from Asia have conducted serosurveys for COVID-19 in their general population. A recently reported community based study from Karachi, Pakistan, has reported a seroprevalence of 8.7% (95% CI 5.1-13.1) and 15.1% (95% CI 9.4-21.7) in low and high transmission areas respectively, with no significant difference between males and females. (30) Similarly, the seroprevalence reported from two community clinics in Tokyo was 3.83% (95% CI 2.76-5.16). (31)While others have reported a higher seroprevalence among the elderly, our study did not find any difference between the age groups with respect to their seropositivity. Thus, the evidence till date is showing significant regional as well as time dependant variations in findings of serosurveys to assess the exposure to SARS-CoV-2.Most serosurveys, including ours have reported a large majority of infections to be asymptomatic in nature. Among the symptomatic cases, the most common symptom was fever followed by cough and diarrhea as a symptom was strongly associated with seropositivity.

An interesting finding is higher seropositivity in members of larger households indicating the higher risk of household transmission among them. Correlation with the actual detected epidemic curves of the three cities show a trend where with higher seroprevalence, there is consistent relative flattening. Thus the possibility of herd immunity being achieved at some point of time in the population cannot be ruled out. GIS analysis shows that wards detected with high seropositivity did not necessarily report more detected cases, implying gaps in testing in those regions.

Our study had a few limitations. The participants were only adults and the non-response rate was high (17.4%), and hence, the possibility of selection bias cannot be excluded. The non response was higher among females probably due to cultural factors and higher individual apprehensions towards blood sample collection. The study reported on the prevalence of antibodies against SARS-CoV-2 at a point in time. Follow up data on anti-SARS-CoV-2 antibodies in the same subjects will be required to understand the duration of immunity to natural infection as well as protection against reinfection. Serial cross-sectional serosurevs have been planned in the same population to address this issue and estimate the rate of spread of COVID-19 infection in Odisha.

To conclude, our study found a high seroprevalence against COVID-19 in urban Odisha. Future studies integrating the seroprevalence data with sociocultural and other biological data will help us better understand the dynamics of COVID-19 transmission and the susceptibility to infection at the individual and community level. It will also help us understand the effectiveness of the several steps undertaken by the state and central Government such as social distancing, usage of masks, etc, in preventing the spread of COVID-19 infection in the community. However, we should be careful while interpreting the findings of a seroprevalence study. There is still no concrete data to support the fact that presence of antibodies against COVID-19 is protective against reinfection. Also, seroprevalence studies should not be used to stigmatize any community or politicized to underestimate the efforts of any government in reducing the spread of infection in their respective countries.

## Data Availability

Data will be made available after peer reviewed publication and on reasonable request to the corresponding author

## Notes

### Competing Interest Statement

The authors have declared no competing interest.

### Funding Statement

Funding for the study has been provided by the Odisha State Government, Department of Health and Family Welfare.

### Author Declarations

RMRC Bhubaneswar Institutional Human Ethical Committee and Odisha State Health Research and Ethics Committee

